# Advancing Drug Development with aiHumanoid Simulations: A Virtual Phase 1 Comparative Study of Standard Chemotherapy versus Standard Chemotherapy plus COTI-2 for Pancreatic Adenocarcinoma

**DOI:** 10.1101/2023.09.08.23295256

**Authors:** WR Danter

## Abstract

**Background:** Traditional Phase 1 trials often provide important drug development insights, which can be limited by ethical issues, costs, and lengthy timelines. Advancements in AI based simulations offer a potential avenue for mitigating these challenges. The present study used the aiHumanoid platform, specifically the upgraded DeepNEU database v8.1, to design and conduct a virtual Phase 1 trial, to assess the comparative efficacy and toxicity of standard chemotherapy with gemcitabine plus Taxol vs a combination of standard chemotherapy plus COTI-2 for treating Pancreatic Adenocarcinoma (PAC).

**Methods:** Applying the updated DeepNEU database of 7267 genotypic and phenotypic concepts linked through 67491 relationships, the study used aiHumanoid simulations to predict outcomes from 30 virtual patients. Data from the standard chemotherapy arm and the standard treatment plus COTI2 arm were analyzed at 25%, 50%, 75%, and 100% of maximal dose. Estimates of efficacy and potential toxicities were based on a combination of the paired 2 tailed T test and Cohen’s d values as a true estimate of treatment effects.

**Results:** The novel combined treatment regimen, especially at 100% dosage, showed medium to large treatment effects on the entire Pancreatic Adenocarcinoma disease profile. Notably, a significant decrease was observed in all disease profile components, bolstered by p-values less than 8.68E-5 and Cohen’s d values >=0.335. While evidence hinted at an increased bone marrow toxicity in the novel treatment arm, no individual organoid toxicity exceeded one standard deviation above predicted values. Importantly, COTI-2 treatment demonstrated a dose-dependent increase in p53 levels, significant at p < 0.006.

**Conclusion:** This aiHumanoid’s virtual Phase 1 trial emphasizes the potential of computational simulations in the drug development process. Our findings indicate a promising treatment pathway combining COTI-2 with standard chemotherapy for Pancreatic Adenocarcinoma. Ongoing development and validation of the aiHumanoid based virtual Phase 1 clinical trial methodology is warranted.

## Introduction

In 2023, PAC remains one of the deadliest cancers worldwide [1]. Despite advances in medical science, late diagnosis, rapid progression, and resistance to conventional treatments make PAC an ongoing public health concern [2, 11]. An intricate molecular and cellular heterogeneity characterizes PAC, complicating the prediction of therapeutic responses [3]. Traditional models, both in vitro and in vivo, while insightful, often fail to capture the complete complexity and dynamic behavior of human disease, particularly in a patient-specific context [5]. Artificial intelligence (AI), however, offers new opportunities to address this challenge [6].

PCA, a malignancy stemming from the pancreatic cells, is notorious for its lethality [1]. Nonspecific symptoms like jaundice, abdominal pain, weight loss, and nausea often make early detection challenging [14]. Diagnosis encompasses blood tests, imaging, and biopsies to establish the presence and magnitude of PAC tumors [14]. Treatment efficacy heavily depends on several factors, including the tumor’s stage, location, molecular characteristics, and the patient’s overall health [15]. However, standard treatments such as surgery, chemotherapy, and radiation therapy, are associated with notable side effects and might not significantly enhance the patient’s survival or quality of life [16].

Recent advances in molecular testing have highlighted PAC’s heterogeneity, revealing different subtypes and mutations that could potentially guide treatment strategies [17]. Some patients, for instance, have inherited mutations like BRCA1/2 or PALB2, rendering them more receptive to specific drugs [17]. Others might have mutations in genes such as KRAS or TP53 that drive the growth and resistance of pancreatic cancer cells [17]. Despite the promise these findings hold, targeted therapies and immunotherapies derived from them are still experimental, and their efficacy remains inconsistent for many PAC patients [17].

The integration of AI in biomedical research presents unprecedented opportunities in disease modeling, drug discovery, and personalized treatments [6]. AI-based simulations, including the emerging aiHumanoid simulations [20], could offer a more nuanced understanding of disease progression and treatment responses [7, 8]. If developed to accurately reflect an individual patient’s physiological behavior and disease progression, such simulations could drastically alter the therapeutic approach to PAC [9]. They could predict the efficacy of innovative drug combinations based on the patient’s response, inching closer to the personalized medicine goal [10]. Nonetheless, aiHumanoid simulations for PAC research are still embryonic, necessitating rigorous efforts for clinical validation and optimization.

While PAC remains a formidable challenge, technological and research advancements, especially the incorporation of AI, offer new hope. Continued efforts to understand the disease’s biology and leverage AI tools might pave the way for improved patient outcomes and a transformative approach to treatment.

In this first of its kind study, we use a series of updated aiHumanoid simulations (v8.1) as virtual patients in a virtual Phase 1 clinical trial to evaluate the impact of adding a small molecule activator of mutant P53, COTI-2 [18,19], to approved first line therapy with Gemcitabine plus Taxol [41,42] for the treatment of PAC. This investigation marks the beginning of our scientific journey to evaluate the potential of replacing human-based Phase 1 drug trials with aiHumanoid-driven simulations.

## Methods

This study employs robust aiHumanoid simulations of PAC and utilizes the literature validated simulations to compare the efficacy and toxicities of conventional chemotherapy with gemcitabine plus Taxol to treatment with the novel combination of gemcitabine plus Taxol plus COTI2, a small molecule activator of mutant p53. The following sections detail the methodology used in this first of its kind virtual Phase 1 trial:

### 1a. Updating the aiHumanoid Simulation to v8.1

The first step in this study involved updating the previous v8.0 of the aiHumanoid [20] to v8.1. The updated version includes new simulations of Prostate, Parathyroid, and Skeletal muscle, increasing the total number of integrated organoid simulations from 18 to 21. The cranial nerve components of the brainstem simulation were updated in v8.1. The whole brain simulated arterial and venous blood supply were also updated, particularly at the capillary level. Regarding literature validation of the simulations, the same approach used in version 8.0 was applied to the new and updated simulations comprising v8.1. A detailed overview of the process is summarized in Appendix 1.

### 1b. PAC Validation Profile in the aiHumanoid Simulation

To confirm the presence of PAC in the aiHumanoid simulation, the following 14 cellular biomarkers features gleaned from the literature were evaluated: CA 19-9, CDKN2A, CEA, desmoplastic reaction, fibroblast activation, KRAS, liquid biopsy, LRG1, Mucin1, Mucin 2, S100A2, S100P, SMAD4, TP53, and TTR [21-27].

The final panel of PAC disease markers was determined by statistical analysis using a Z Score estimate of >2.58 since all patient profile changes from control were in a positive direction. The final panel of predictors for the healthy virtual patient profile and 30 virtual PAC patients were compared first with a correlation coefficient and a corrected p Value <0.006 to determine whether PAC patient profiles were indeed significantly different from the healthy control.

The series of 30 virtual PAC patient profiles was created by GPT4 from recent literature to cover a wide range of patient demographics, phenotypic and genomic characteristics than could be achieved with a single common patient simulation. All patient profiles include Age, Gender, Risk factors, Symptoms, Tumor characteristics, Mutation profile and CA 19-9 levels. A representative group of virtual patient profiles is summarized in Appendix 2.

### 2. Rationale for drugs and drug combinations

The validated aiHumanoid PAC simulation was used to evaluate the response of each of the 30 virtual patients to one of two drug combinations. The current standard of care for advanced PAC is generally one of two options. The first, Folfirinox, a combination of Folinic acid, 5FU, Irinotecan and Oxaliplatin, is effective in high functioning patients with PAC [43]. The second approved combination contains gemcitabine plus the Nab form of Taxol (paclitaxel) and is used in patients judged unlikely to tolerate Folfirinox. These two forms of chemotherapy are approved in multiple countries including Canada, the US and EU member states.

Approximately 50-70% of PAC are associated with a p53 loss of function mutation. These mutations make PAC more difficult to treat effectively with current first line chemotherapy options. Compounds that could even partially restore mutant p53 activity could be a definite advancement in the treatment of PAC. COTI-2 is a novel third generation thiosemicarbazone that has successfully completed Phase 1 human trials in gynecological cancers. Importantly COTI-2 has both p53 dependent and independent pathway effects. Research has demonstrated the ability of COTI-2 to refold mutant p53 to a more normal configuration with at least partial restoration of normal function. In addition, COTI-2 appears to have modulating effects on the PI3K-AKT-mTOR pathways. The completed Phase 1 trial with COTI-2 established an acceptable safety profile and a Phase 2 dose (https://www.clinicaltrials.gov/study/NCT02433626). An early efficacy signal was noted in the trial that included heavily pretreated participants. Finally, preclinical work demonstrated in vitro synergy with several chemotherapeutic agents including Gemcitabine and Taxol in pancreatic cancer cells [18].

### 3. Study Design and Objectives

This is the first of its kind, open-label virtual Phase 1 clinical trial. The dual primary objectives are to determine the safety and early-stage efficacy of COTI-2 when combined with the standard of care (Gemcitabine + (Nab)Taxol) in patients with PAC.

The virtual patients: The profiles for 30 unique and virtual patients diagnosed with PAC were synthesized by GPT-4, OpenAI’s advanced language model (July 2023 version) https://chat.openai.com/. GPT4 used its extensive training data encompassing medical literature, patient profiles, and related clinical information, to synthesize diverse representative patient backgrounds, histories, genetic markers, and disease stages. Each virtual patient was crafted to reflect a broad spectrum of demographic characteristics, risk factors, and clinical manifestations associated with PAC, ensuring a comprehensive representation for the purpose of our study. The 30 virtual patients used in this study serve as hypothetical but commonly encountered examples but do not represent real individuals or precise medical histories.

#### Inclusion criteria for virtual patients

1. Algorithmically confirmed diagnosis of PAC, using an optimized profile of cellular markers associated with confirmed PAC.
2. All virtual patients were generated as treatment-naive or having received simulated prior therapies that did not include previous treatment with gemcitabine, Taxol, or COTI-2.
3. Virtual patients included male and female patients aged 50 years or older.
4. Adequate simulated organoid health including cardiac, bone marrow, liver, lung, kidney and neurological function. The health statuses of 21 organoids are represented as predicted data points within an acceptable range for the virtual wild type or unaffected aiHumanoid.

#### Exclusion criteria for virtual patients

1. Simulated treatment with any other experimental agents or anti-cancer therapies prior to or concurrent with study enrollment.
2. Virtual patients with a prior history of gemcitabine, Taxol, or Coti-2 exposure.
3. History of virtual allergic reactions attributed to compounds of similar composition to gemcitabine, Taxol, or COTI-2.
4. A history of uncontrolled virtual cardiovascular disease.
5. Virtual patients with active simulated CNS metastasis unless their status has been stable without symptoms for 6 months prior to enrollment.
6. Uncontrolled concurrent virtual illnesses, including but not limited to, ongoing or active digital infections or psychiatric conditions.
7. Virtual pregnant or breastfeeding patients.
8. Virtual patients with a history of other malignancies within the 2 virtual years prior to enrollment

##### Treatment and Dose Escalation

###### Dosing

Given the unique nature of virtual patients, it is possible to use all 30 patients in each of the two arms of the study. In Arm 1, the virtual patients will receive increasing doses of gemcitabine plus (Nab)Taxol. In Arm 2, all patients will receive Gemcitabine plus (Nab)Taxol plus oral COTI-2. The specific doses for both arms will be first 25%, then 50%, next 75%, and finally 100% of maximal dose. Dose escalation from 25% through 100% percent of maximal dose will continue after the safety and preliminary efficacy of the previous dose level have been determined.

###### Safety Assessments

The safety of conventional and novel treatment is assessed using organoid based cellular toxicity markers derived from the current literature. An optimized subset of 11 organoid toxicity markers was used to evaluate toxicity of both treatment arms in all virtual patients. These organoid specific markers can be found in Appendix 3.

For the purposes of this study dose-limiting organoid specific toxicities (DLTs) are defined compared to the conventional therapy with gemcitabine plus Taxol. Here we define a DLT as any individual patient data point in any of the toxicity markers that is more than 2 standard deviations above the mean for the 30 virtual patients that received gemcitabine plus Taxol.

### 4. Assessment of Response to Therapy Using a Specific Biomarker Subset

An optimal composite biomarker profile subset derived from the original list of 11 features was used to assess the response to therapy in the simulated PAC patients. Changes in the levels of these biomarkers post-treatment with the gemcitabine + Taxol + COTI2 arm, relative to their gemcitabine + Taxol arm, serve as indicators of therapeutic efficacy.

#### Simulation Analysis and Interpretation

Step 1: For evaluating efficacy and toxicity differences between treatment arms, a paired two tailed T test was employed with a required p value of 0.006 for statistical significance.

Step 2: To further evaluate differences between treatment arms, Cohen’s d [28,29] was used to estimate real treatment effects as small, medium, or large.

Step 3: In the event of discordant estimates between p values and Cohen’s d values, decisions were based on the Cohen’s d value.

Step 4: Within group differences were conducted in an analogous manner except that:

Step 5: To assess individual patient treatment associated toxicity, a Z-test was used to evaluate where each of the 30 virtual patients from Arm 2 (gemcitabine + Taxol + COTI-2) stood in the distribution of toxicities seen in Arm1 (gemcitabine + Taxol). Any Arm 2 patient toxicity estimate >= 2 SD above the mean of the Arm 1 toxicity estimates was defined as a DLT.

### 5. Statistical Analysis

Null hypothesis: The novel combination of COTI-2 with gemcitabine and paclitaxel (Taxol) does not result in any difference in treatment outcomes for virtual PAC patients when compared to the standard of care (gemcitabine and (Nab)Taxol). Given that multiple comparisons being conducted, a Bonferroni correction was applied, making the adjusted p value for rejecting the null hypothesis p < 0.006. The alternate hypothesis states that the combination of gemcitabine, Taxol and COTI2 produces better treatment outcomes for virtual PAC patients when compared to the standard of care (gemcitabine and (Nab)Taxol).

To assess the response to therapy, levels of an optimized subset of biomarkers (see Results section) were measured both before and after treatment interventions in both arms. The two-tailed paired t-test was conducted for each biomarker to determine if there was a statistically significant difference (p<0.006) in their levels between pre-treatment post treatment results. In addition, Cohen’s d was calculated to estimate the true treatment effects.

## Results

In the present study, v8.1 (2023) of the DeepNEU database was used, which was characterized by several upgrades in contrast to its most recent previous version v8.0, [20]. Firstly, v8.1 consisted of 7267 genotypic and phenotypic concepts that were linked through 67491 positive and negative concepts. This indicates that for every nonzero concept in the relationship matrix, there were more than 9.4 incoming and outgoing relationships.

Furthermore, v8.1 offered new validated simulations for Prostate, Parathyroid, Skeletal muscle, and cranial nerves as part of the brainstem simulation, increasing the total number of integrated organoid simulations from 18 to 21. The whole brain simulated arterial and venous blood supply were also expanded. The previously implemented improved early stopping algorithm (v8.0) was retained in this version, proving effective as the multiple networks consistently converged, achieving optimal results after 14-17 iterations. For the purposes of this research, we chose to use the results after 16 iterations, as determined through a 3 valued moving average, which avoided noticeable overfitting after this point.

### Virtual patient demographics

The gender specific characteristics of the 30 virtual PAC patients are summarized in Table 1. The data indicate that the patients are well matched. There were no statistically significant differences between male and female attributes at the p = 0.05 level. Importantly, all 30 virtual patients were treatment naïve at the time of enrolment.

**Table 1.**

| Virtual Patient Factor | Men | Women | Total | p value |
| --- | --- | --- | --- | --- |
| Gender | 15 (50.0) | 15 (50.0) | 30 | NS |
| Age (yrs) | 63±7 | 58±5 | 61±4 | NS |
| P53 mutations | 9 (64.3) | 5 (35.7) | 14 | NS |
| KRAS mutation | 8 (47.1) | 9 (52.9) | 17 | NS |
| CDKN2A mutation | 5 (45.5) | 6 (54.5) | 11 | NS |
| SMAD4 | 8 (72.7) | 3 (27.3) | 11 | NS |
| Treatment Naïve | 15 (50.0) | 15 (50.0) | 30 | NS |
| CA 19-9 expression* | 0.854±0.028** | 0.821±0.106** | 0.838±0.066** | NS |
| * range 0 to +1 |  |  |  |  |
| ** ± 95% confidence interval |  |  |  |  |

### The Pancreatic cancer feature profile optimization

Data for the 30 virtual patients was generated and analyzed for predictive value by comparing the profile with that from an unaffected aiHumanoid. A Z test was used to identify which subset of the 14 original features best predicted the presence of PAC A two tailed paired T test was then used to confirm the predictive value of the final profile. The original p value of 0.05 was modified for multiple comparisons using the Bonferroni correction. This process created a corrected p value of 0.006. Using this method 8 of the original 14 features were selected for inclusion in the final disease profile. These eight factors are CA 19-9, CEA, Desmoplastic reaction, Fibroblast activation, LRG1, Mucin 2, S100P and TTR. A summary of this analysis appears in Table 2

**Table 2:** Final feature profile for confirming the presence of PAC.

| Virtual Patient | CA 19-9 | CEA | Desmoplasia | Fibroblast activation | LRG1 | Muc2 | S100P | TTR |
| --- | --- | --- | --- | --- | --- | --- | --- | --- |
| No PAC | -1 | -1 | -0.067 | -0.881 | -0.9 | -0.54 | -0.4807 | 0.1288 |
| PAC | 0.52801 | 0.67531 | 0.740163 | 0.35499 | 0.906077 | 0.473297 | 0.471423 | 0.80118 |
|  |  |  |  |  |  |  | *p = | 7.95E-05 |

\*p value from a 2 tailed paired T test, Bonferroni corrected $p < 0.006$ needed to reject the null hypothesis that the feature profile is no different between no PAC and PAC virtual patients

### Dose Escalation

#### Dose 1: 25% of Maximum Dose

The initial phase of formal testing started with a dose equivalent to 25% of the maximal dose for all drugs in both treatment arms. We utilized Cohen’s d to estimate the treatment effect. A two-tailed paired T-test was also used to determine the significance of the predicted differences observed between the gemcitabine plus Taxol arm and the gemcitabine plus Taxol plus COTI-2 arm. While T-test p-values provided insight, our primary metric for discerning true treatment effects was the Cohen’s d estimate, as recommended by sources including [28] and [29].

For assessing toxicity, our analysis mirrored the above approach, but a Z-score was added to gauge treatment toxicities in arm 1 in comparison to the predicted toxicities for arm 2. A Z-score of >=2 was set as a benchmark to identify potential toxic outliers in the second arm. Using this approach, any treated patient with a Z-score of >=2 was considered to have experienced a dose-limiting toxicity.

**Table 3a:**
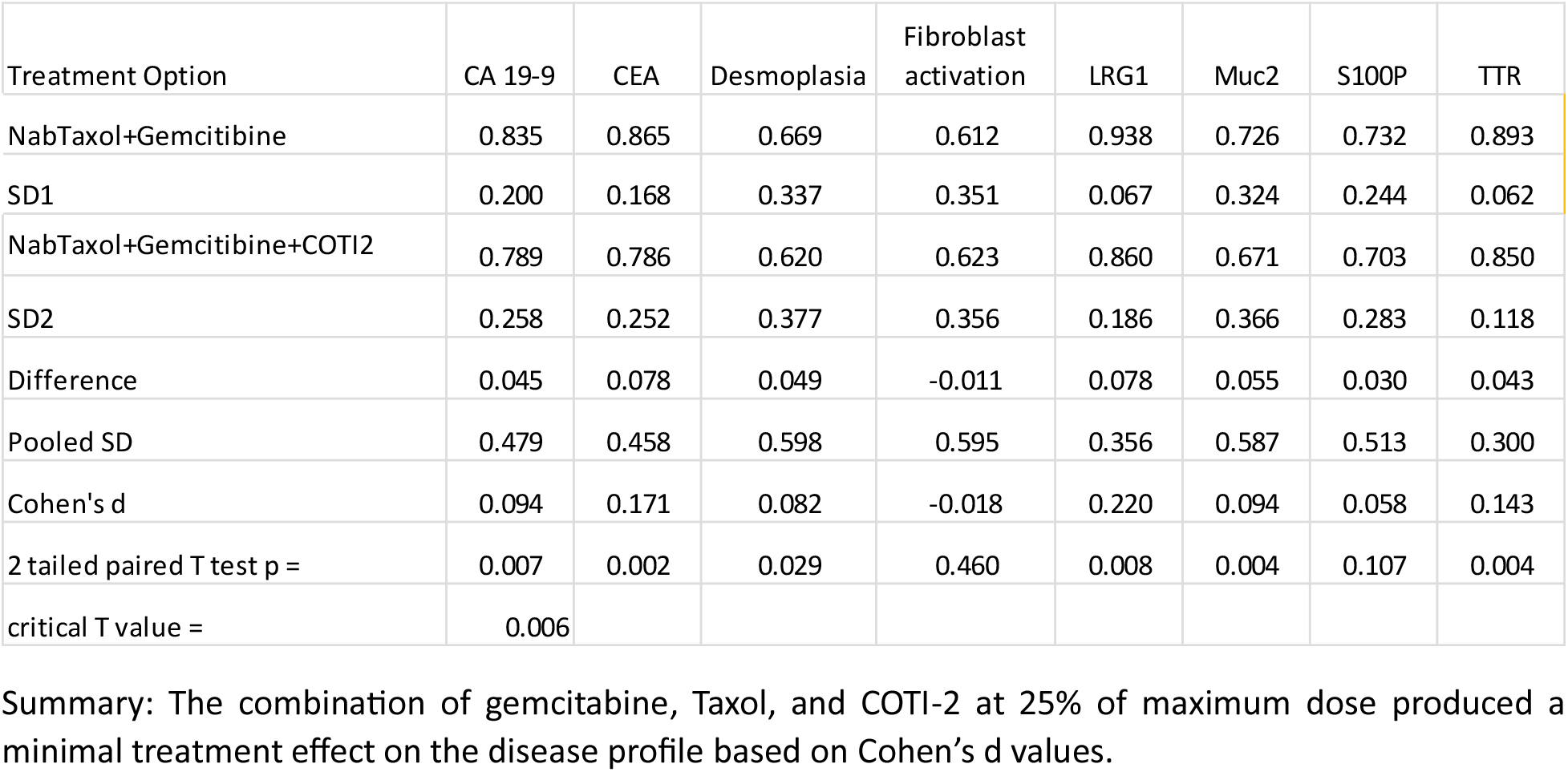
Efficacy at 25% of maximum dose.

**Table 3b:**
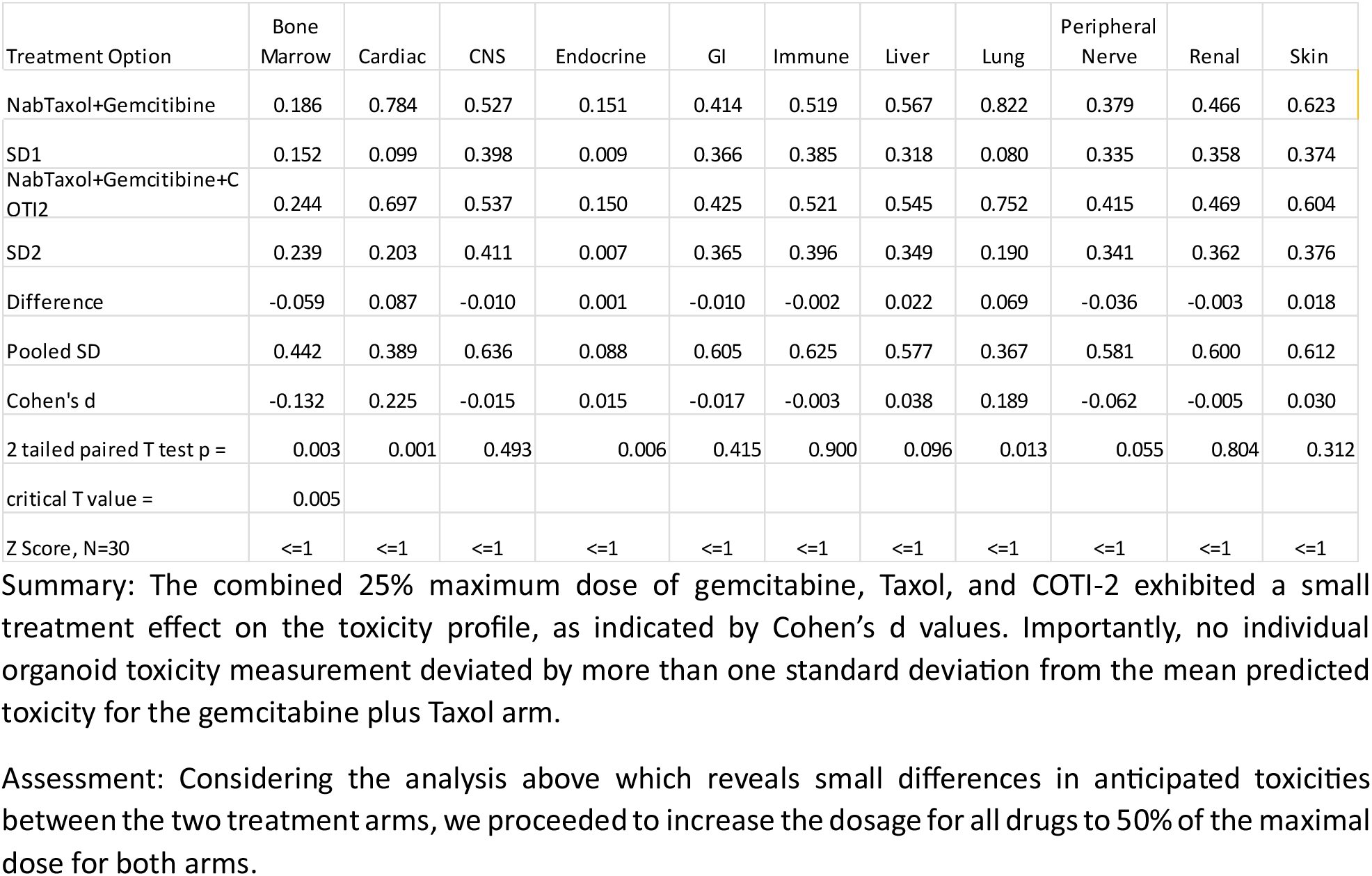
Organoid specific toxicities at 25% of maximum dose.

#### Dose 2: 50% of maximum dose

The analysis of all patient data was conducted in an identical manner to that outlined above for Dose 1 Efficacy and Toxicity.

**Table 4a:** Efficacy at 50% of maximum dose.

| Treatment Option | CA 19-9 | CEA | Desmoplasia | Fibroblast activation | LRG1 | Muc2 | S100P | TTR |
| --- | --- | --- | --- | --- | --- | --- | --- | --- |
| NabTaxol+Gemcitabine | 0.846 | 0.853 | 0.659 | 0.628 | 0.936 | 0.703 | 0.739 | 0.886 |
| SD1 | 0.197 | 0.178 | 0.338 | 0.355 | 0.079 | 0.338 | 0.246 | 0.072 |
| NabTaxol+Gemcitabine+COTI2 | 0.663 | 0.652 | 0.579 | 0.611 | 0.683 | 0.602 | 0.579 | 0.696 |
| SD2 | 0.370 | 0.375 | 0.379 | 0.375 | 0.374 | 0.388 | 0.388 | 0.286 |
| Difference | 0.184 | 0.201 | 0.080 | 0.017 | 0.254 | 0.101 | 0.160 | 0.190 |
| Pooled SD | 0.533 | 0.526 | 0.599 | 0.604 | 0.476 | 0.603 | 0.563 | 0.423 |
| Cohen's d | 0.345 | 0.382 | 0.134 | 0.028 | 0.532 | 0.167 | 0.284 | 0.449 |
| 2 tailed paired T test p = | 4.74E-05 | 0.000128 | 0.004513849 | 0.014154721 | 0.000211 | 0.000882 | 4.11E-05 | 0.000108 |
| critical T value = | 0.00625 |  |  |  |  |  |  |  |

Summary: The combination of gemcitabine plus Taxol and COTI-2 at 50% of maximum dose produced a modest treatment effect on several components of the PAC disease profile when compared with the gemcitabine plus Taxol arm based on Cohen's d values. The greatest impact was seen in significant decrease in LRG1 value while the least effect was seen for the Fibroblast activation prediction.

**Table 4b:** Organoid specific toxicities at 50% of maximum dose.

| Treatment Option | Bone Marrow | Cardiac | CNS | Endocrine | GI | Immune | Liver | Lung | Peripheral Nerve | Renal | Skin |
| --- | --- | --- | --- | --- | --- | --- | --- | --- | --- | --- | --- |
| NabTaxol+Gemcitabine | 0.302 | 0.702 | 0.576 | 0.202 | 0.459 | 0.554 | 0.576 | 0.723 | 0.470 | 0.496 | 0.625 |
| SD1 | 0.159 | 0.099 | 0.406 | 0.014 | 0.365 | 0.394 | 0.320 | 0.083 | 0.337 | 0.359 | 0.376 |
| NabTaxol+Gemcitabine+COTI2 | 0.334 | 0.562 | 0.548 | 0.165 | 0.419 | 0.507 | 0.513 | 0.570 | 0.459 | 0.446 | 0.577 |
| SD2 | 0.323 | 0.322 | 0.415 | 0.030 | 0.348 | 0.393 | 0.370 | 0.353 | 0.328 | 0.361 | 0.385 |
| Difference | -0.032 | 0.139 | 0.028 | 0.038 | 0.041 | 0.046 | 0.063 | 0.152 | 0.011 | 0.051 | 0.048 |
| Pooled SD | 0.491 | 0.458 | 0.641 | 0.148 | 0.597 | 0.627 | 0.587 | 0.467 | 0.577 | 0.600 | 0.617 |
| Cohen's d | -0.065 | 0.304 | 0.044 | 0.254 | 0.068 | 0.074 | 0.107 | 0.326 | 0.019 | 0.084 | 0.078 |
| 2 tailed paired T test p = | 0.000 | 0.000 | 0.332 | 0.098 | 0.105 | 0.005 | 0.000 | 0.000 | 0.000 | 0.002 | 0.112 |
| critical T value = | 0.005 |  |  |  |  |  |  |  |  |  |  |
| Z Score, N=30 | <=1 | <=1 | <=1 | <=1 | <=1 | <=1 | <=1 | <=1 | <=1 | <=1 | <=1 |

#### Dose 3: 75% of maximum dose

**Table 5a:**
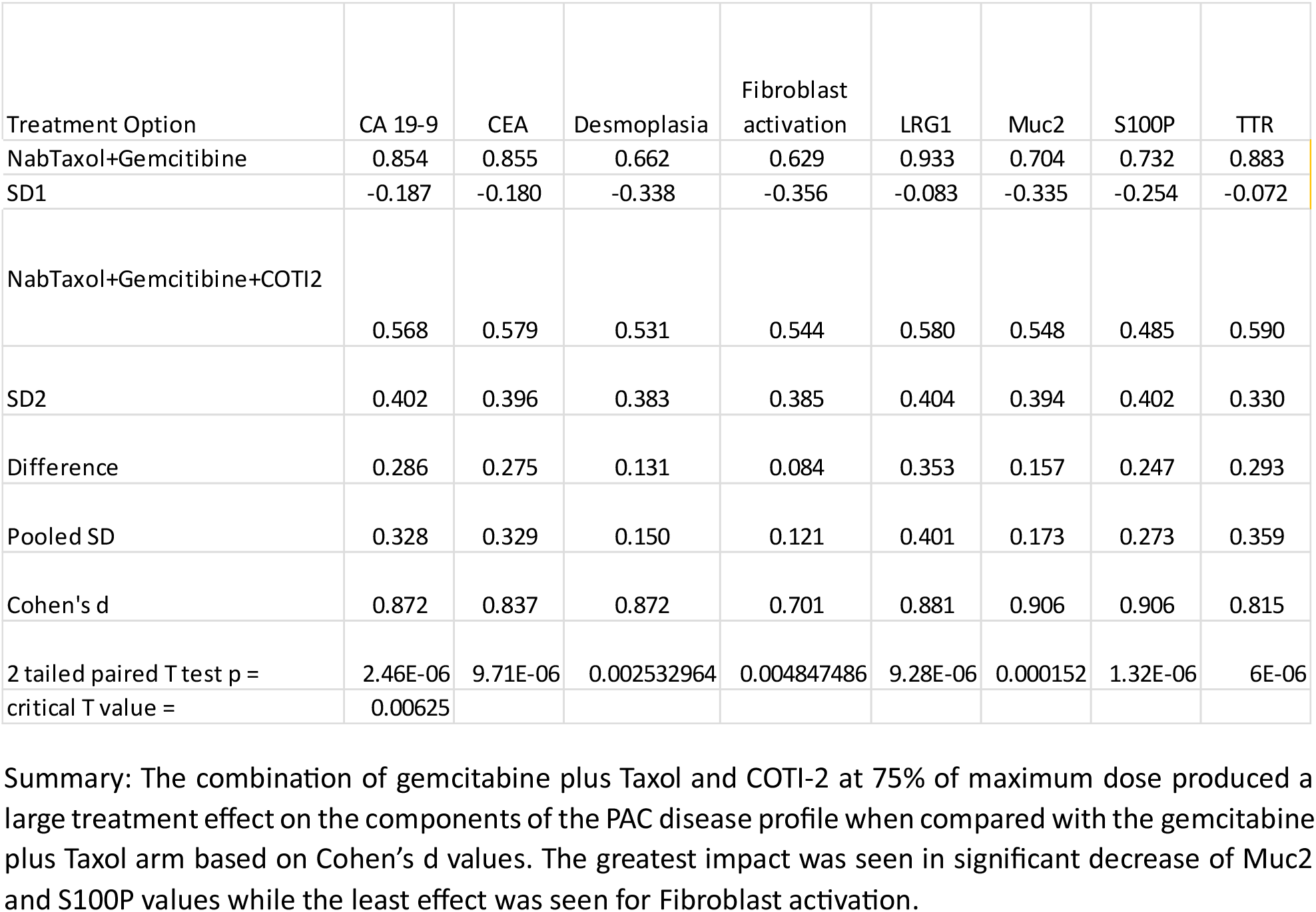
Efficacy at 75% of maximum dose.

**Table 5b:**
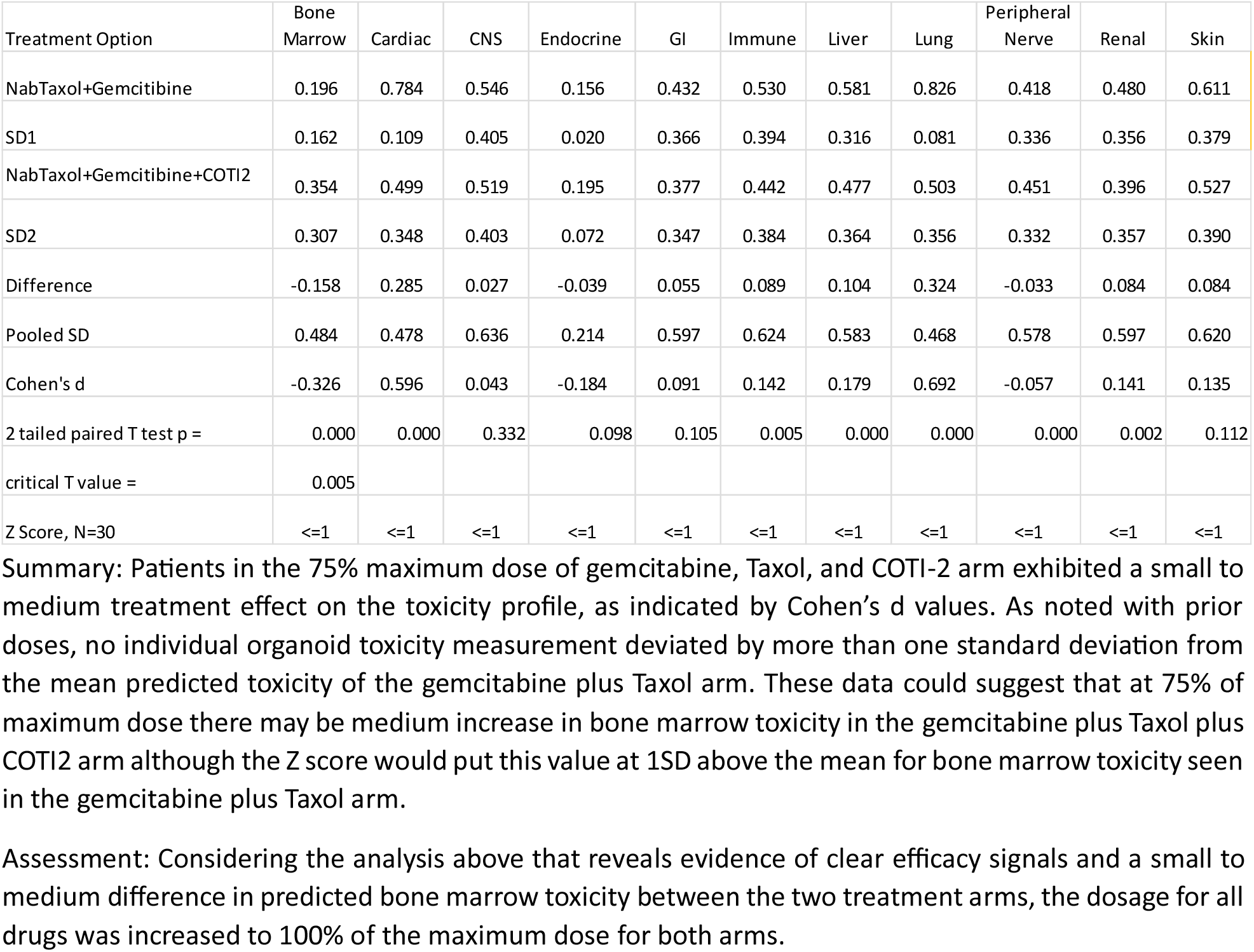
Organoid specific toxicities at 75% of maximum dose.

#### Dose 4: 100% of maximum dose

**Table 6a:** Efficacy at 100% of maximum dose.

| Treatment Option | CA 19-9 | CEA | Desmoplasia | Fibroblast activation | LRG1 | Muc2 | S100P | TTR |
| --- | --- | --- | --- | --- | --- | --- | --- | --- |
| NabTaxol+Gemcitabine | 0.849 | 0.852 | 0.661 | 0.639 | 0.943 | 0.706 | 0.734 | 0.875 |
| SD1 | 0.196 | 0.179 | 0.339 | 0.360 | 0.059 | 0.334 | 0.249 | 0.083 |
| NabTaxol+Gemcitabine+COTI2 | 0.417 | 0.436 | 0.409 | 0.439 | 0.461 | 0.438 | 0.376 | 0.469 |
| SD2 | 0.404 | 0.390 | 0.364 | 0.355 | 0.397 | 0.366 | 0.382 | 0.298 |
| Difference | 0.432 | 0.416 | 0.252 | 0.200 | 0.482 | 0.268 | 0.359 | 0.406 |
| Pooled SD | 0.548 | 0.534 | 0.593 | 0.598 | 0.477 | 0.591 | 0.562 | 0.437 |
| Cohen's d | 0.789 | 0.779 | 0.425 | 0.335 | 1.010 | 0.453 | 0.639 | 0.931 |
| 2 tailed paired T test p = | 5.05E-08 | 8.68E-05 | 2.9043E-05 | 6.97218E-08 | 3.48E-06 | 2.19E-08 | 2.37E-09 | 3.04E-08 |
| critical p value = | 0.00625 |  |  |  |  |  |  |  |

Summary: The combination of gemcitabine plus Taxol and COTI-2 at 100% of maximum dose produced a medium to large treatment effect on all components of the PAC disease profile when compared with the gemcitabine plus Taxol arm based on Cohen's d values. A statistically significant decrease in all components of the PAC disease profile was observed with all 2 tailed p values less than 8.68E-5 and all Cohen's d values $\geq 0.335$ .

**Table 5b:**
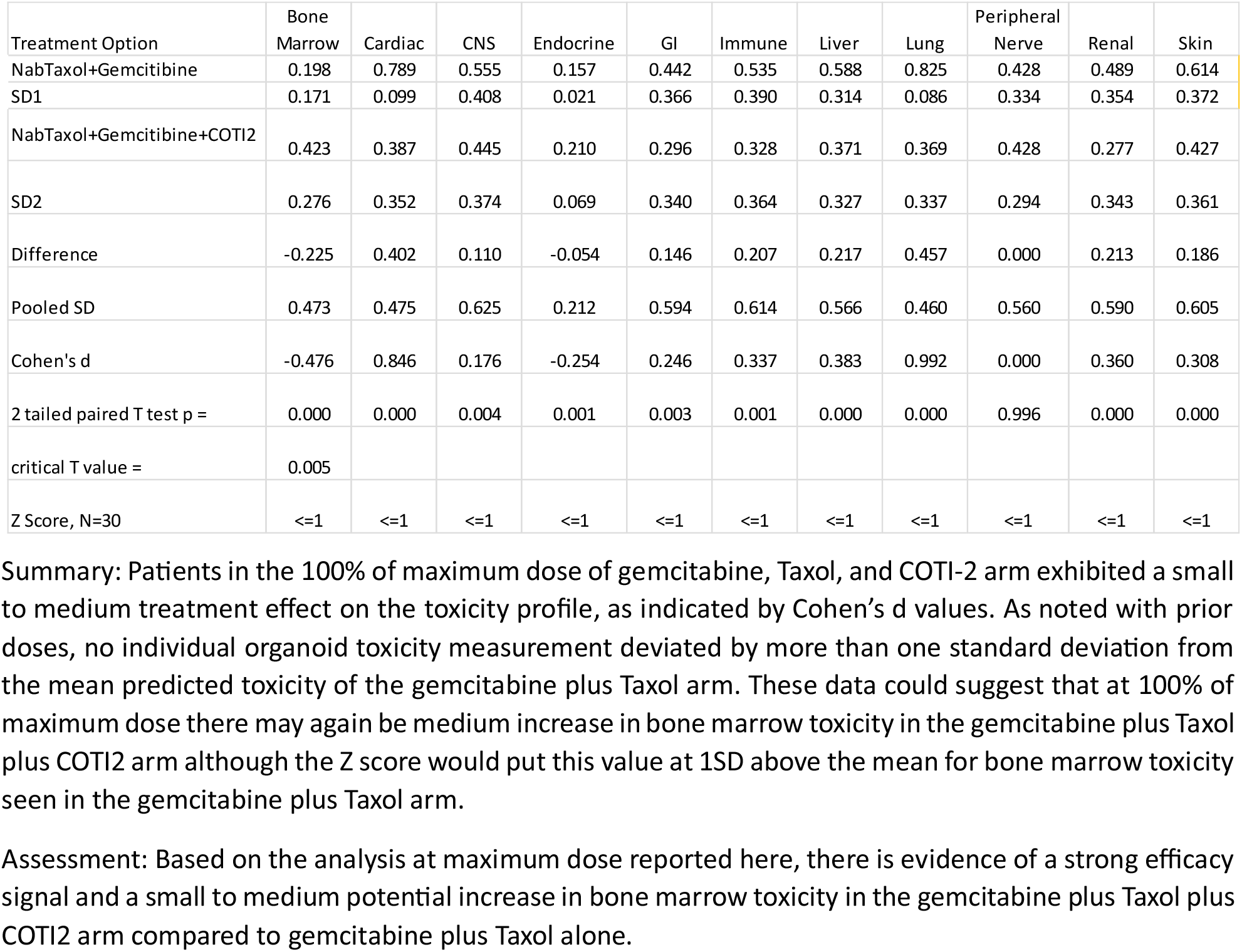
Organoid specific toxicities at 100% of maximum dose.

#### COTI2 and p53 levels

Treatment with COTI2 produced a dose dependent increase in treatment effect on p53 levels. Based on Cohen’s d values, the treatment effect begins at small and increases to a medium-large effect. All treatment two tailed p values were significant at the p < 0.006 level.

**Table 7:** Dose response effects on P53 levels.

| Treatment Option | p53-25% | p53-50% | p53-75% | p53-100% |
| --- | --- | --- | --- | --- |
| NabTaxol+Gemcitabine | 0.122 | 0.117 | 0.115 | 0.115 |
| SD1 | 0.266 | 0.254 | 0.248 | 0.248 |
| NabTaxol+Gemcitabine+COTI2 | 0.207 | 0.306 | 0.394 | 0.448 |
| SD2 | 0.268 | 0.263 | 0.248 | 0.241 |
| Difference | 0.085 | 0.189 | 0.280 | 0.334 |
| Pooled SD | 0.517 | 0.509 | 0.498 | 0.494 |
| Cohen's d | 0.165 | 0.371 | 0.562 | 0.675 |
| 2 tailed paired T test p = | 5.05E-08 | 8.68E-05 | 2.9E-05 | 6.97E-08 |
| critical p value = | 0.006 |  |  |  |

Note- p53-25% refers to the p53 activity at 25% of maximum dose etc

## Discussion

The present study represents a significant advancement in the application of the DeepNEU platform v8.1 to assess potential treatment options for pancreatic adenocarcinoma (PAC). This version of the platform represents a substantial upgrade over its predecessor [20], integrating a broad array of genotypic and phenotypic concepts that pave the way for more realistic simulations. The updates in the underlying database have allowed for simulations that capture nuances of the relationships between organoids, with the notable addition of organoid simulations including Prostate, Parathyroid, Skeletal muscle, and the Brainstem (cranial nerves) integrated with a more robust CNS blood supply simulation.

The analysis of the PAC feature profile, based on data from 30 virtual patients, revealed eight crucial factors including CA 19-9, CEA, Desmoplastic reaction, Fibroblast activation, LRG1, Mucin 2, S100P, and TTR that optimally predict the presence of PAC. These factors, verified through rigorous statistical processes, could provide clinicians with a robust framework for detecting and treating PAC in its initial stages. The efficacy and toxicity profiles of the combination of gemcitabine, Taxol, and COTI-2, established at various dosage levels, offer a glimpse into the potential therapeutic benefits and associated risks of the novel treatment regime in the context of a large unmet need.

A significant point of debate raised by this study is the revolutionary potential of aiHumanoid simulations to replace traditional Phase 1 trials. Traditional Phase 1 trials, while central to drug development, have been plagued by challenges such as prolonged durations, high costs [30], ethical concerns surrounding human trials [31], and limited generalizability due to smaller sample sizes [32]. There is also the aspect of predictive power; traditional models do not always accurately forecast human responses due to inherent differences [33].

Addressing these concerns, the aiHumanoid Phase 1 virtual clinical trial represents a potentially transformative solution. By enabling comprehensive AI based computational trials, these simulations offer improved efficiency, reduced costs, and bypass the ethical dilemmas associated with animal testing and early human trials. Moreover, with the capability to simulate a broader range of genetic and physiological diversity, they provide a more holistic testing environment. One cannot overlook the potential for enhanced predictive power afforded by these simulations either, as they meticulously consider countless variables spanning molecular, cellular, and systemic levels.

However, the path to replacing traditional Phase 1 trials with aiHumanoid simulations is unlikely to be simple. Although the FDA has recently signaled that it may eventually be open to nontraditional approaches like computer simulations [34], rigorous scientific development and real-world validation will be necessary to ensure the accuracy and reliability before any widespread adoption of AI driven virtual Phase 1 clinical trials. As aiHumanoid technology evolves, incorporating comprehensive feedback mechanisms and aligning them closely with real-world outcomes will be critical.

It’s clear that the combination of cutting-edge technologies, like artificial intelligence (AI), and medical research holds the key to a future where drug development is safer, more inclusive, and remarkably efficient. The present research demonstrates that we’re on the cusp of a transformative era in medical science, and it’s imperative to continue this trajectory based on a rigorous scientific method.

Finally, based on the results from this virtual Phase 1 trial demonstrating increased efficacy with modest potential for increased bone marrow toxicity, an actual Phase1b/2 trial with gemcitabine plus Taxol with COTI2 in Pancreatic adenocarcinoma is warranted.

## Limitations and Future Directions

The introduction of aiHumanoid simulations into the field of PAC research marks an important step forward in the domain, despite the inherent challenges. A primary limitation of this technology stems from the reliance on the quality and diversity of data employed to train these models. The richer the dataset, encompassing a wide array of factors such as genetics, epigenetics, and proteomics, the more accurate and personalized the simulations become [35]. Thus, the major hurdle lies in gathering such comprehensive data from a diverse patient population. At present the DeepNEU database covers ∼32% of the human genome. It is our goal to reach ∼99% coverage in 3 years’ time.

Secondly, the current aiHumanoid model has demonstrated significant promise in identifying a potential drug combination, yet there is an urgent need to enhance the model’s accuracy in forecasting therapeutic responses [36]. This is crucial in mitigating the risk of false-positive results, which could lead to unnecessary and potentially burdensome clinical trials.

A third limitation is the extensive computational requirements associated with these aiHumanoid simulations. As models become increasingly sophisticated and based on larger genomic datasets, the need for computational power escalates. It therefore becomes a priority to continually optimize the algorithm’s efficiency without compromising its predictive power [6].

Looking forward, the innovative field of aiHumanoid simulations presents significant potential to revolutionize PAC treatment. To actualize this potential, future research should concentrate on refining these models and expanding their abilities. This includes integrating added dimensions of data, such as information about the microbiome, and further incorporating physiological and biological processes [37].

As we progress towards an era of personalized medicine, it becomes imperative for aiHumanoid simulations to become more tailored to individual patients. This involves generating models that consider a patient’s genetic, proteomic, and clinical profile in much greater detail. In doing so, aiHumanoid simulations can offer even more precise and personalized treatment options [38].

Furthermore, efforts should be made to make these complex aiHumanoid simulations more accessible for healthcare providers. While the underlying technology may be intricate, its results should be interpretable and applicable by clinicians, thus ensuring effective implementation in patient care [39].

Building on the promise shown in cancer research, aiHumanoid simulations and virtual clinical trials also hold the potential to revolutionize our approach to other medical conditions, most notably in the field of rare diseases.

### Beyond cancer virtual trials: Rare diseases

Virtual clinical trials would also represent a profound change for rare diseases research, where traditional clinical trials often face substantial hurdles in patient recruitment and availability. Due to the scarcity of patients suffering from rare conditions, assembling a sufficiently large sample size for a conventional clinical trial can be time-consuming, costly, and sometimes impractical. For example, a sufficiently rare disease trial could require 50 centers around the globe to recruit a small number of patients. Virtual trials, which leverage remote monitoring technologies and data collection methods, offer a way to overcome these limitations. They enable patients to participate from the comfort of their own homes, irrespective of their geographic location. This broadens the recruitment pool and makes it feasible to collect adequate data for statistical validation. By reducing the need for frequent hospital visits, virtual trials also make participation less burdensome for patients, who may already be dealing with debilitating symptoms. Overall, the flexibility and accessibility of virtual clinical trials could accelerate the pace of rare disease research, expediting the development and approval of much-needed treatments.

In summary, aiHumanoid simulations represent a groundbreaking extension to the toolkit of cancer and rare disease research [40]. While challenges remain, their potential for advancing personalized medicine is both compelling and transformational.

## Data Availability

All data produced in the present work are contained in the manuscript

## Appendix 1 Technology Overview: aiHumanoid Simulation using Artificial Intelligence

### 1. Introduction

The aiHumanoid project uses the DeepNEU Artificial Intelligence (AI) platform to create highly realistic computer simulations of human organoids. Grounded in pioneering stem cell research, the project aims to replicate human organoid systems with computer simulations. It incorporates complex growth media and integrates individual organoids through a common cardiovascular system into a holistic aiHumanoid simulation.

### 2. Methodology

i. Simulation of Induced Pluripotent Stem Cells (aiPSCs): The project begins with the AI-driven simulation of induced pluripotent stem cells (aiPSCs), from simulated human fibroblasts. These simulated cells are exposed to the 4 Yamanaka transcription factors to start growth and development, mimicking human stem cell behavior.
ii. Growth Media Development: A single, unified growth medium suitable for all 21 organoid types was developed. It combines DMEM/F12, B27 supplement, and Matrigel. Organoid-specific media were also created by adding to this base formula, to offer more accurate simulations for specific organoids such as bone marrow, liver, and lungs etc.
iii. Creation of Wild-Type Organoids: After validation, aiPSCs were used to derive several types of organoids. Two approaches are employed: an unguided pathway for neural organoids and a guided pathway, involving specialized media, for non-neural organoids.
iv. Integration into aiHumanoid: All individual organoids are integrated with a simulated cardiovascular system for realistic nutrient supply, oxygenation, and waste removal. This overcomes the limitations associated with in vitro organoid systems based on diffusion which leads to restricted growth and central necrosis.
v. Validation and Statistical Analysis: Organoid simulations are validated using an extensive set of cell specific markers derived from the peer reviewed literature. Statistical analysis was done using a two-tailed Z-test due to lack of standard deviation information.

### 3. Scientific Rigor

The model incorporates statistical analysis to evaluate the null hypothesis concerning the differences in expression levels between male and female aiHumanoids, and between different conditions like uninfected and gram-negative sepsis simulations.

### 4. Implications and Future Work

The aiHumanoid project has successfully simulated a variety of organoids and integrated them into a functional system, advancing the use of AI in biomedical research. These simulations have broad applications, ranging from pharmaceutical development to disease modeling.

This novel technology offers a highly practical alternative to wet lab and animal experiments for simulating human physiology and sets the stage for further refinements and potential real-world applications in medicine and research.

## Appendix 2 Representative virtual patient profiles used in this virtual Phase 1 trial

### Patient A Early Detection, Familial Risk

- Age: 50 years
- Gender: Female
- Risk factors: Strong family history of pancreatic cancer, BRCA2 mutation
- Symptoms: Detected through regular screening due to high risk; asymptomatic
- Tumor characteristics: Localized to pancreas, no metastasis
- Mutation profile: BRCA2 mutation, CDKN2A mutation
- CA 19-9 level: Slightly elevated

### Patient B Typical Presentation

- Age: 70 years
- Gender: Male
- Risk factors: Smoker, chronic pancreatitis
- Symptoms: Abdominal pain, jaundice, weight loss
- Tumor characteristics: Tumor in head of pancreas, local lymph node involvement
- Mutation profile: KRAS, TP53, CDKN2A, and SMAD4 mutations
- CA 19-9 level: Significantly elevated

### Patient C Advanced Disease at Presentation

- Age: 75 years
- Gender: Female
- Risk factors: Diabetes, obesity
- Symptoms: Back pain, jaundice, weight loss, nausea
- Tumor characteristics: large tumor in body of pancreas, liver metastasis
- Mutation profile: KRAS, TP53, CDKN2A, and SMAD4 mutations
- CA 19-9 level: Significantly elevated

### Patient D Late Stage, High Performance Status

- Age: 65 years
- Gender: Male
- Risk factors: Smoker, heavy alcohol use
- Symptoms: Fatigue, abdominal pain, weight loss
- Tumor characteristics: Widespread metastasis to liver and lungs
- Mutation profile: KRAS, TP53, and CDKN2A mutations
- CA 19-9 level: Very high

### Patient E Rare Genetic Profile

- Age: 60 years
- Gender: Female
- Risk factors: Family history of breast cancer, previous breast cancer diagnosis
- Symptoms: Jaundice, steatorrhea (fat in stool)
- Tumor characteristics: Tumor in tail of pancreas, local lymph node involvement
- Mutation profile: ERBB2 (HER2) amplification
- CA 19-9 level: Elevated

## Appendix 3 Summary of Drug Induced Toxicity markers

### Bone Marrow Toxicity

CD34+, CD117 (c-Kit), Hemoglobin levels, White Blood Cell Count (WBC)

### Cardiac Toxicity

Troponin I and T, Brain Natriuretic Peptide (BNP), B-type Natriuretic Peptide

### Central Nervous System (CNS) Toxicity

Glial Fibrillary Acidic Protein (GFAP), Neuron-Specific Enolase (NSE), S100 Calcium-Binding Protein B (S100B)

### Endocrine Toxicity

Thyroid-Stimulating Hormone (TSH), Insulin, Cortisol

### Gastrointestinal (GI) Toxicity

Intestinal Fatty Acid-Binding Protein (I-FABP), Citrulline, Gastrin

### Immune System Toxicity

CD3 (T cells), CD19 (B cells), CD56 (NK cells)

### Liver Toxicity

Alanine Aminotransferase (ALT), Aspartate Aminotransferase (AST), Alkaline Phosphatase (ALP), Bilirubin

### Lung Toxicity

Surfactant Protein D (SP-D), Krebs von den Lungen-6 (KL-6), Clara Cell Protein 16 (CC16)

### Peripheral Nerve Toxicity

Neurofilament Light (NFL), Myelin Basic Protein (MBP), Ganglioside GM1 Antibodies

### Renal Toxicity

Kidney Injury Molecule-1 (KIM-1), Cystatin C, Neutrophil Gelatinase-Associated Lipocalin (NGAL), Beta-2-microglobulin

### Skin Toxicity

Epidermal Growth Factor Receptor (EGFR), Melanoma Antigen Recognized by T cells 1 (MART-1), Cytokeratin

## Acknowledgement

The author acknowledges and is grateful to Dr Linda Pullan for her expert review and recommended edits that improved the quality of this manuscript.

